# Factors associated with peri-neonatal mortality in Sao Tome & Principe: a prospective cohort study

**DOI:** 10.1101/2022.08.07.22278476

**Authors:** Alexandra Vasconcelos, Swasilanne Sousa, Nelson Bandeira, Marta Alves, Ana Luísa Papoila, Filomena Pereira, Maria Céu Machado

**Affiliations:** Unidade de Clínica Tropical - Global Health and Tropical Medicine (GHTM), Instituto de Higiene e Medicina Tropical (IHMT), Universidade NOVA de Lisboa. Lisboa - Portugal; Maternidade, Hospital Dr. Ayres de Menezes, São Tomé, República Democrática de São Tomé e Príncipe; CEAUL, NOVA Medical School/Faculdade de Ciências Médicas, Universidade NOVA de Lisboa, Lisboa - Portugal; Professora Catedrática Jubilada da Faculdade de Medicina de Lisboa, Universidade de Lisboa. Lisboa – Portugal

**Keywords:** neonatal mortality, perinatal mortality, stillbirth, neonatal death, Sao Tome & Principe

## Abstract

**Background:** Neonatal mortality reduction is a global goal but its factors in high-burden nations vary and are poorly understood. This study was conducted to identify the factors affecting peri- neonatal mortality in Sao Tome & Principe (STP), one of the smallest African countries.

**Methods:** Institution-based prospective cohort study conducted at Hospital Dr. Ayres Menezes. Maternal-neonate dyads enrolled were followed up after the 28^th^ day of life (n=194) for identification of neonatal death-outcome (n=22) and alive-outcome groups (n=172). Data were collected from pregnancy cards, hospital records and face-to-face interviews. After the 28^th^ day of birth, a phone call was made to evaluate the newborn’s health status. A logistic regression model was used to identify the relevant factors associated with mortality, a p value <0.05 was considered statistically significant.

**Results:** The mean gestational age of the death-outcome and alive-outcome groups was 36 (SD=4.8) and 39 (SD=1.4) weeks, respectively. Death-outcome group (n=22) included sixteen stillbirths, four early and two late neonatal deaths. High-risk pregnancy score [cOR 2.91, 95%CI:1.18-7.22], meconium-stained fluid [cOR 4.38, 95%CI:1.74–10.98], prolonged rupture of membranes [cOR 4.84, 95%CI:1.47–15.93], transfer from another unit [cOR 6.08, 95%CI:1.95–18.90], and instrumental vaginal delivery [cOR 8.90, 95%CI:1.68–47.21], were factors significantly associated with mortality. The odds of experiencing death were higher for newborns with infectious risk, IUGR, resuscitation maneuvers, fetal distress at birth, birth asphyxia, and unit care admission. Female newborn [cOR 0.37, 95%CI:0.14-1.00] and birth weight of more than 2500 g [cOR 0.017, 95%CI:0.002-0.162] were found to be protective factors. In the multivariable model, meconium-stained fluid was significantly associated with death outcome.

**Conclusion:** Factors such as having a high-risk pregnancy score, meconium-stained amniotic fluid, prolonged rupture of membranes, being transferred from another unit, and an instrumental- assisted vaginal delivery increased 4– to 9–fold the risk of stillbirth and neonatal death. Of the factors associated with peri-neonatal mortality in this study, avoiding health-worker-related factors associated with delays in prompt intrapartum care is a key strategy to implement in Sao Tome & Principe.

## Introduction

The first 28 days of life—the neonatal period—are the most vulnerable time for a child’s survival, and a significant number die before birth never having the chance to take their first breath. Perinatal mortality includes stillbirths and early neonatal deaths (ENND), indicating the death of a live newborn before the age of seven completed days [1]. Late neonatal deaths (LNNDs) are those that occur after 7 days to 28 completed days of birth [2].

Globally, perinatal mortality accounts for three-fourths of deaths during the neonatal period [3,4]. More than half of the cases of stillbirths occur when pregnant women are in labor, and these deaths are directly related to the lack of skilled care at this critical time [5,6]. On the other hand, the largest contributors to neonatal mortality (ENND plus LNND) are complications of preterm birth, birth asphyxia, infection, and congenital malformations although they can differ depending on the country context [7].

Only in the last two decades, mainly after Lawn et al published article titled “4 million neonatal deaths: When? Where? Why?” [8], attention started to be given to the neonatal period in developing countries although stillbirths are still invisible and missing from the Sustainable Development Goals (SDG) agenda [5,9,10]. It urges to highlight that stillbirths and newborn deaths account for twice as many deaths as malaria and human immunodeficiency virus (HIV) infection combined but have received much less awareness and funding in these resource-constrained countries [8,9].

Understanding the causes of stillbirths (fetal deaths) is also complex, as there are many promoting and interacting factors [11]. In most low –to medium - income countries (LMICs), it is difficult to determine the exact reason for the stillbirth; therefore, the cause of death is often classified as “unexplained” [5]. Researchers report that different risk factors for fetal death, such as maternal factors (advanced maternal age, high pre-pregnancy body mass index, smoking, low socioeconomic status), obstetric history (grand multiparity, previous stillbirth), antepartum factors (fewer than four antenatal visits, fetal growth restriction, maternal anemia, maternal fever and infections, antepartum hemorrhage, hypertension), and intrapartum factors (preterm birth, extremes of neonatal birth weight, cesarean delivery, operative vaginal delivery, and assisted breech delivery), are all factors that have been reported in various studies as causes of stillbirths [6,11–15].

Sao Tome & Principe (STP) is an LMIC with low HIV/AIDS prevalence and a malaria preelimination phase [16,17]. There is an established antenatal care (ANC) service, with a 98% rate of women seen at least once by a skilled health provider during pregnancy and a 95.4% rate of deliveries occurring in health units [17]. In STP, peri-neonatal mortality is considered a public health problem in the country, given that neonatal deaths accounts for approximately 43% of all under5 deaths [18]. At the time this study was initiated, there was an annual rate of 22 stillbirths and 22 newborn deaths per 1000 livebirths [18].

We are aware that the concept of knowing what works in terms of reducing perinatal and neonatal mortality is complicated by a huge diversity of country contexts and of determinants of maternal and neonatal health [19–22]. However, according to Lawn and other authors, identifying and addressing avoidable causes of neonatal death is possible even in poorly functioning health systems, justifying our current study [8,9]. Answering - why, when, and where - newborns die in Sao Tome & Principe will enable the design of appropriate planning to prevent this major public health problem since, for appropriate prevention of fetal and newborn mortality, data pertaining to its determinants are important.

To the best of our knowledge, this is the first study to assess peri-neonatal mortality in STP, and it was undertaken within the context of a broader project on neonatal adverse birth outcomes (ABOs) in this LMIC [23–25].

Therefore, we conducted a prospective cohort study to identify the most important factors associated with mortality among newborns delivered at HAM.

## Materials and methods

### Study design

An institution-based prospective cohort study was conducted at Hospital Dr. Ayres de Menezes (HAM) for mother-neonate dyads followed up until the 28^th^ day after delivery (neonatal period).

### Setting

The archipelago of Sao Tome & Principe is one of the smallest sub-Saharan African (SSA) countries, with approximately 200.000 inhabitants and a total land surface of approximately 1,001 km^2^ on two islands (Sao Tome and a smaller island named Principe) [16,17]. The rural districts have ANC services and maternity units with basic obstetric services in Lembá, Lobata, Caué, Me-Zochi, and Principe Island, except for the district of Cantagalo. There is only one hospital in the country, Hospital Dr. Ayres de Menezes (HAM), with the only maternity in the country with Comprehensive Emergency Obstetric and Neonatal Care (EmONC) capable of providing blood transfusions and performing cesarean sections [26]. As a tertiary healthcare facility, HAM receives the most complicated cases from facilities in lower levels of care. There is no health insurance policy in the country or any private maternity units. In STP, there are approximately 3.2 doctors per 10 000 inhabitants and 27 nurses per 10 000 inhabitants, and at HAM, there are three to four obstetricians and one to two general doctors who provide care to the neonates [26]. There are no neonatologists in the country. The midwives in the labor ward are responsible for the initial resuscitation of normal deliveries, and doctors from the pediatric department are called to the labor ward to attend babies in distress.

The HAM maternity unit has a facility-based clinical care unit for ill newborn babies, but there is no neonatal or child intensive care unit in the country. The Neonatal Care Unit (NCU) receives high-risk babies delivered within the institution and referrals from other health facilities or from home with a total capacity to admit six babies. This unit, like others in SSA settings, is basic and able to manage simple neonatal complications such as hypothermia, feeding problems and sepsis suspicion. Although NCU was rebuilt in 2016, there is still a lack of continuous positive airway pressure therapy, surfactant therapy and enteric feeding for assisting sick babies.

### Study population and follow-up

All mother-neonate dyads admitted to the HAM maternity unit for childbirth constituted the source population whereas the study populations were selected neonates delivered in the HAM maternity unit during the study period. Recruitment of participants (mother-newborn dyad) occurred from July 2016 to November 2018. During the study period, 4540 deliveries were recorded, corresponding to 450 cesarean deliveries and 3740 normal vaginal births.

The inclusion criteria for participants were as follows: 1) all neonates delivered at HAM with a gestational age of 28 weeks or more and 2) newborns who were born outside the hospital but were later admitted at HAM. A total of 537 newborns were initially enrolled.

The exclusion criteria included the following: 1) all neonates delivered at HAM with a gestational age of less than 28 weeks, 2) newborns whose mothers had no antenatal pregnancy card, 3) newborns whose mothers had cognitive impairment. The newborn was also excluded if his health status was unknown at his twenty-eight days of life.

Consenting participants in the sample were followed up (mother-newborn dyad) throughout their stays until hospital discharge. The survival status of neonates after discharge was ascertained by making a follow-up mobile phone call at the end of the neonatal period. Those who could not be reached by phone after four attempts in different weeks were taken as nonrespondents, and the mother-newborn dyad was excluded. A flowchart of participation in the study is shown in Fig 1.

**Figure 1.**
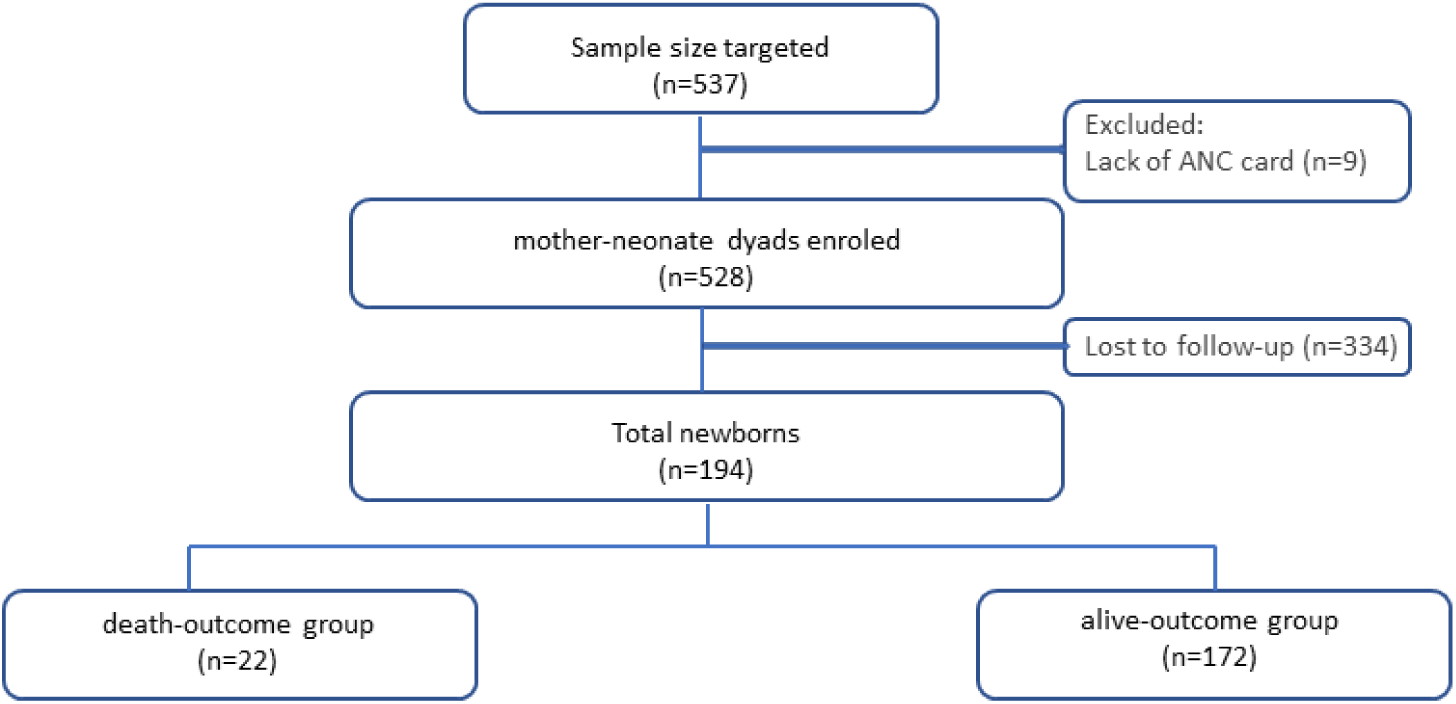
Flowchart of participation in the study

### Sampling method

This is a substudy undertaken to estimate adverse birth outcomes (ABOs) and related risk factors in the country.

The sample size initially calculated for the broader project, using Raosoft® (http://www.raosoft.com/samplesize.html), was based on the formula for sample size and margin of error from the software, reaching the minimum sample size of S□=□355, with 355 (95%) and 579 (99%) interval confidence. The original study included 537 mother-neonate dyads enrolled based on the following assumptions: two-sided 95% confidence level, power of 80% to detect an odds ratio of at least two for ABO. Since the sample size was not calculated for present outcomes, to assess peri-neonatal mortality, a power analysis was performed, varying from 77% to 87% for outcomes such as gestational age (GA<37) for this study. In this study, 194 mother-neonate dyads were included. Participants were selected through random sampling. Each morning, from the pile of motherś medical folders, every second interval folder was selected and then carried on requesting contentment for enrollment. To guarantee a sample with few biases and effects by means of confounding variables, the study was conducted in different months (two weeks every two other months), avoiding seasonal interference (rain season and malaria period). Women were interviewed only after the delivery, although the invitation and consent were obtained during her admission to the maternity unit before birth (live birth or stillbirth).

### Selection of groups: death and alive neonatal outcome

A total of 194 mother-neonate dyads were included and were divided into two groups according to the newborn status (alive versus dead) at the end of the neonatal period. Newborns who died before the 28^th^ day of life (death-outcome group with stillbirths included; n=22) were compared with newborns alive at the 28^th^ day of life (alive-outcome group; n=172) concerning maternal, antepartum, and intrapartum characteristics.

### Study Variables

The dependent variables for the study were death (stillbirth and neonatal - early and late - death) and live newborns.

The definition of stillbirth used was the WHO/ICD (for international comparison and reporting) as a baby born without any signs of life at or after 28 weeks of gestation or at least 1000 g in birth weight. [9,27]. Stillbirths were also described according to whether they were an intrapartum or antepartum stillbirth. Intrapartum stillbirth was defined as a dead-born fetus where intrauterine death occurred after the onset of labor and before birth (fresh stillbirth) [9]. Antepartum stillbirth is a dead born fetus where intrauterine death occurs before the onset of labor (macerated stillbirth) [9].

Neonatal mortality was defined as deaths in the first 28 days of life among live births. Death within the first 24 h (<24 h) or first day of life was recorded in units of completed minutes or hours of life [2]. Deaths occurring ≥24 h of life were recorded in days, from day 1 to 27 completed days [2]. The neonatal death definition used in this study was the one from the World Health Organization (WHO) as “deaths among live births during the first 28 completed days of life” [1], which were further subdivided into early neonatal deaths (ENND) as deaths between 0 and 7 completed days of birth and late neonatal deaths (LNND) as deaths after 7 days to 28 completed days of birth [2,28].

For neonates who died at home after discharge, probable causes of death were assigned by the principal investigator (pediatrician) using the International Statistical Classification of Diseases and Related Health Problems ICD-11 coding. For neonates who died in the HAM, the cause of death was assigned by physicians, who work in the hospital, and confirmed by the principal investigator.

The independent variables tested in this study included factors grouped into five categories: 1) newborns’ maternal sociodemographic factors (age, educational status, occupation, marital status, partner’s education, and residence); 2) preconception factors (previous contraceptive utilization), plus current obstetric condition (gravidity, parity, previous abortion and stillbirth, previous cesarean section and preceding birth interval); 3) ANC service (number of visits, gestational age at first ANC visit, obstetric ultrasound, number of fetuses in the ultrasound) plus antepartum factors as positive ANC screenings (high-pregnancy risk score, maternal anemia as hemoglobin concentration <11 g/dl, bacteriuria, hyperglycemia, Rh incompatibility, intestinal parasitic infection (IPI), malaria, HIV, syphilis, hepatitis B virus and sickle cell); 4) health facility-related factors (being transferred from another unit, who assisted the delivery and partograph use) plus intrapartum factors as mode of delivery and complications (fetal malpresentation [29], umbilical cord complication [30], prolonged rupture of membranes [31,32], meconium-stained amniotic fluid [33], postpartum hemorrhage as bleeding >500 mL, preeclampsia defined as hypertension ≥140/90 mmHg and proteinuria in dipsticks in women who were normotensive at ANC, and obstructed labor [34]); 5) newborns characteristics as (gestational age, sex, birth weight) and complications (intrauterine growth restriction (IUGR), congenital anomalies, infectious risk, neonatal resuscitation, fetal distress at birth, birth asphyxia and admission at NCU) [35–37].

### Data collection

Data on antepartum, intrapartum, and postpartum characteristics of participants were gathered and collected from ANC pregnancy cards, obstetric maternal and newborn clinical records. For antepartum data, relevant details of the perinatal history and antenatal period were collected systematically from the ANC pregnancy card. Intrapartum data were collected from labor follow-up sheets, delivery summaries and maternal medical records. Postpartum data were abstracted from newborn birth charts and/or newborn medical records if admitted to the NCU.

Maternal sociodemographic characteristics were supplemented with a structured administered questionnaire through a face-to-face interview of the mothers 12-24 hours after delivery, similar to other studies from LMICs [38].

### Data quality alive-outcome group

The questionnaires were administered in Portuguese, the country national language. The questionnaire was pretested at HAM one month before data collection in 23 mothers, and modification was made based on the pretest result, mainly adjusting terminology for more culturally friendly terms. Womeńs consent to participate in the study was obtained at the time of admission at HAM, but the interview was held after a woman was stabilized and ready to be discharged. Continuous follow-up and supervision of data collection were made by the supervisors. The collected data were checked daily for completeness. The principal investigator (pediatrician) executed and was responsible for all main activities as follows: 1) obtaining consent and enrollment of the participants, 2) data collection from antenatal cards plus maternal clinical and newborns’ records, 3) newborns’ clinical observation (for diagnosis confirmation), 4) face-to-face interviews, 5) administering all phone interviews and 6) data collection entry into the database.

### Data management

The anonymity and safety of the participants were ensured. Data were secured in a confidential and private location. Participants were referred to by identification numbers, and the informed consent forms were kept separate from the questionnaires. Both could only be linked by a coding sheet available only to the principal investigator.

### Data analysis

Data were entered into the QuickTapSurvey app (©2010-2021 Formstack), and the dataset was exported to Excel for cleaning and further analysis using the Statistical Package for the Social Sciences for Windows, version 25.0 (IBM Corp. Released 2017. IBM SPSS Statistics for Windows, Version 25.0. Armonk, NY: IBM Corp.). All data were checked for completeness and accuracy by the principal investigator and a qualified biostatistician.

Descriptive statistics, namely, frequencies and percentages were estimated. A univariable analysis was also performed. In this study, the neonatal death-outcome group was coded as 1, and the neonatal alive-outcome group was coded as 0. The proportion of missing data ranged from 0.8 to 10% across variables, and missing values higher than 10% were described in the analysis.

Univariable logistic regression analysis was used to estimate the unadjusted effect of each independent variable on the dependent variable. Variables that had a p value less than 0.25 were considered in the multivariable model. A p value < 0.05 was considered statistically significant in multivariable logistic regression analysis.

### Ethics approval and consent to participate

The Ministry of Health of Sao Tome & Principe and the main board of the Hospital Dr. Ayres de Menezes are dedicated ethics oversight bodies, and both approved this study. At the time of the study protocol was submitted (year 2016), there were no ethics committees in STP. All methods were performed in accordance with the relevant guidelines and regulations in practice. Written informed consent was obtained from all participants (or their parent or legal guardian in the case of adolescent under 16 or for illiterate pregnant women) after the purpose of the research was explained orally by the principal investigator. Participants’ or their legal representatives’ also consented to have the results of this research work published. Participation in the survey was voluntary, as participants could decline to participate at any time during the study.

## Results

A total of 194 newborns were followed up during their first 28 days of life. In this study, the newborn’s mean gestational age (GA) was 38.86 weeks with a standard deviation (SD) of 2.26 (minimum 28 - maximum 42 weeks). The death-outcome group (n=22) had a mean GA and birth weight of 36 (SD=4.83) and 2515 (SD=997) g, respectively, while their counterparts (172 in the alive-outcome group) had 39 (SD=1.41) weeks and 3209 (SD=507) g, respectively. The mean maternal age was 27.14 years, with a SD of 6.86 (minimum 15 - maximum 43) years old. The mean maternal age for the death-outcome group and alive- outcome group was 30.73 (SD=7.45) and 26.68 (SD=6.66) years, respectively.

The death-outcome group under-study included 16 stillbirths (72.7%), four (18%) ENND and two (9%) LNND. Stillbirths were 69% intrapartum stillbirths (fresh stillbirths), and 31% were antepartum stillbirths (macerated stillbirths). Stillbirth characteristics are further described in Additional file 1, and the early and late neonatal deaths (ENND and LNND) are described in Additional file 2.

The maternal characteristics as well as antepartum, intrapartum, and postpartum factors for the total of the participants and for the death-outcome group versus the alive-outcome group are described in Tables 1 and 2.

**Table 1.**
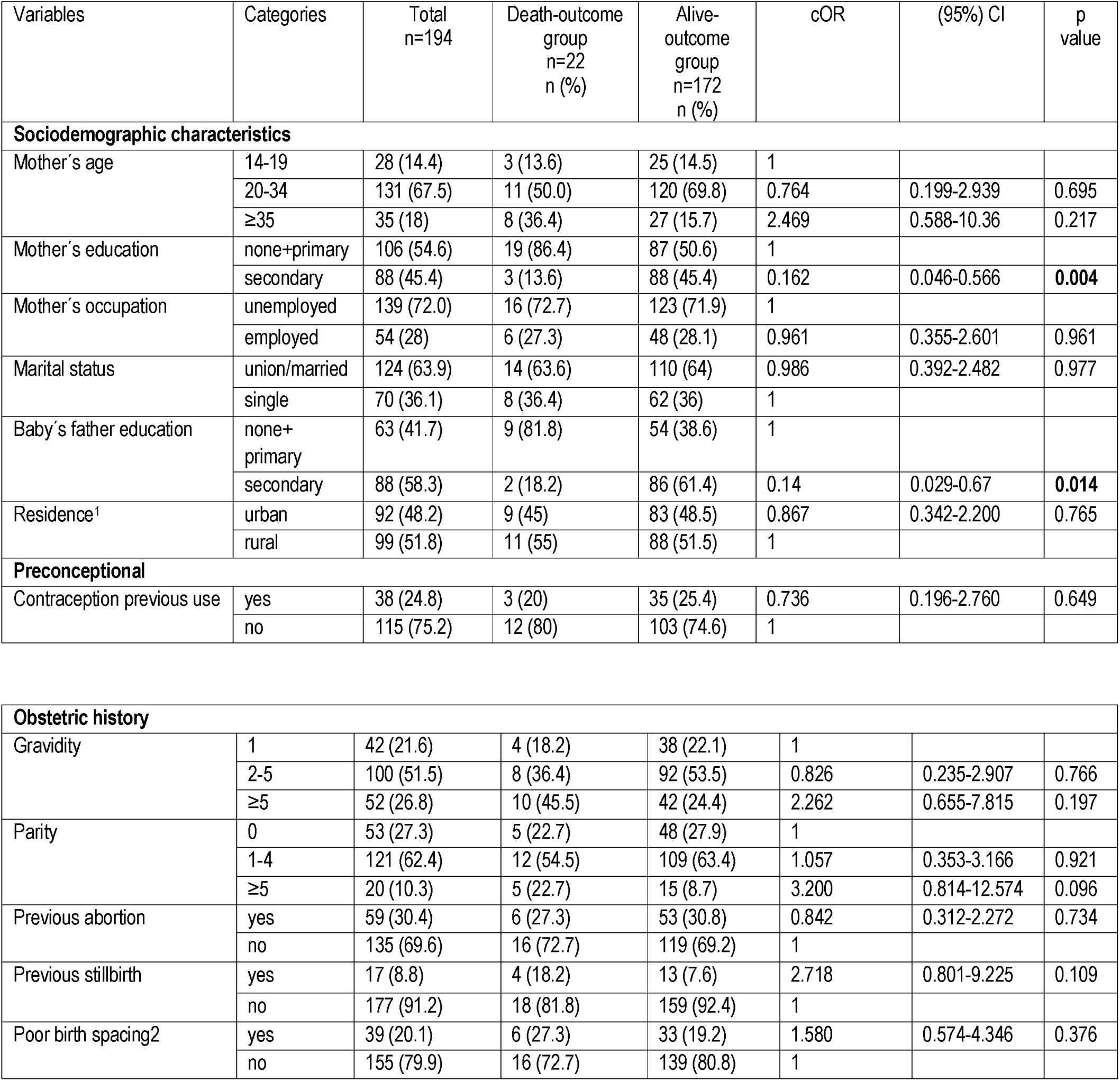
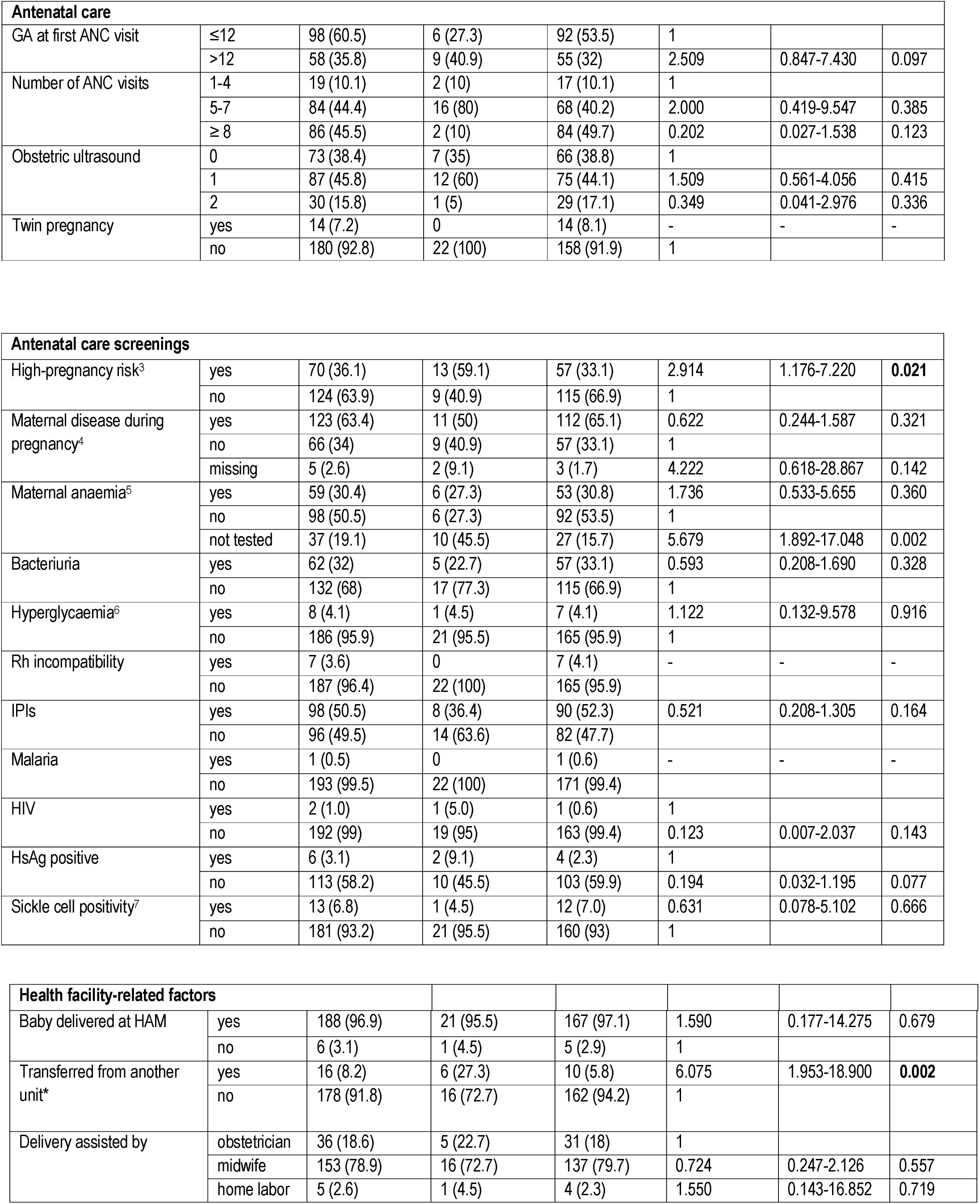
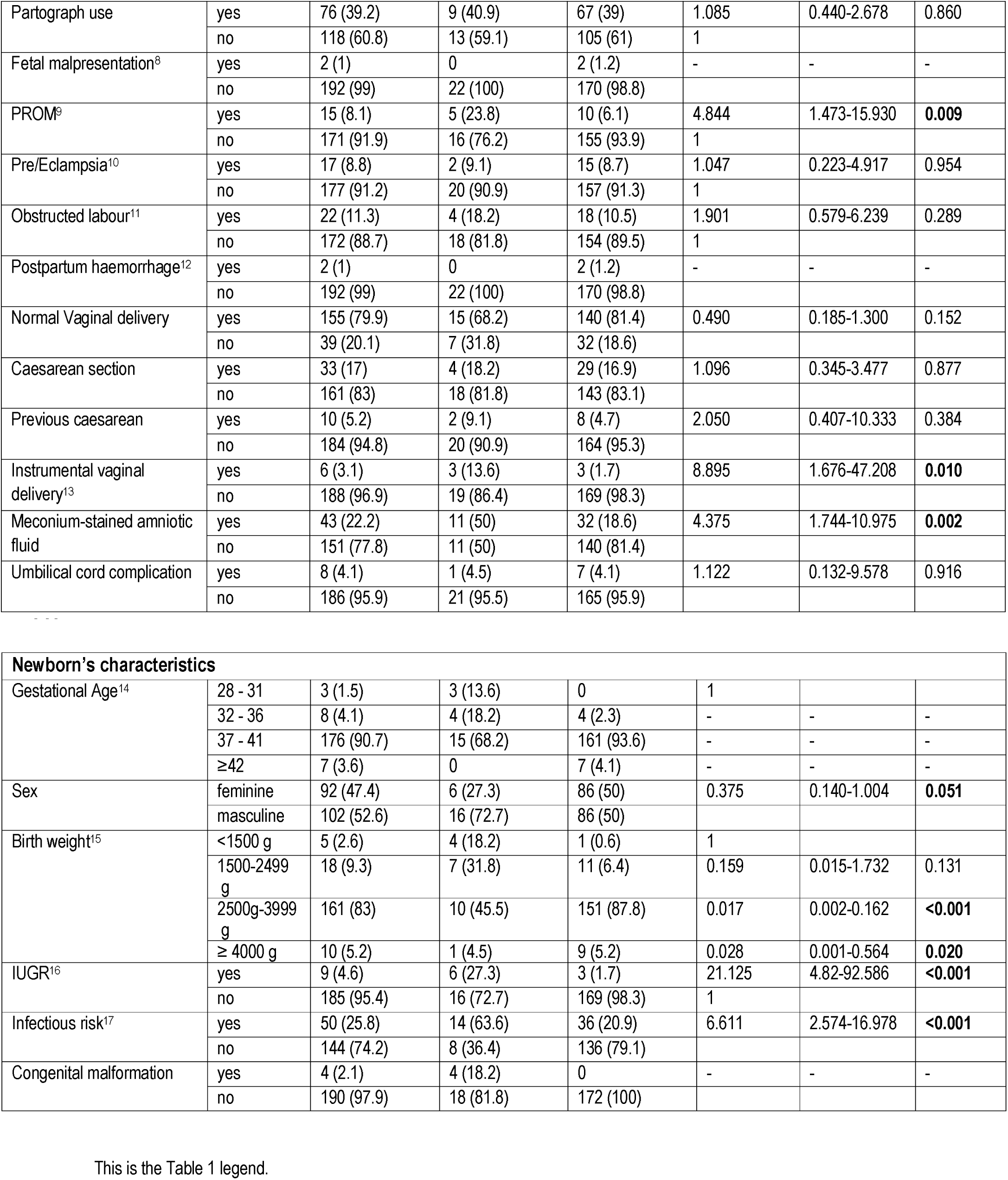
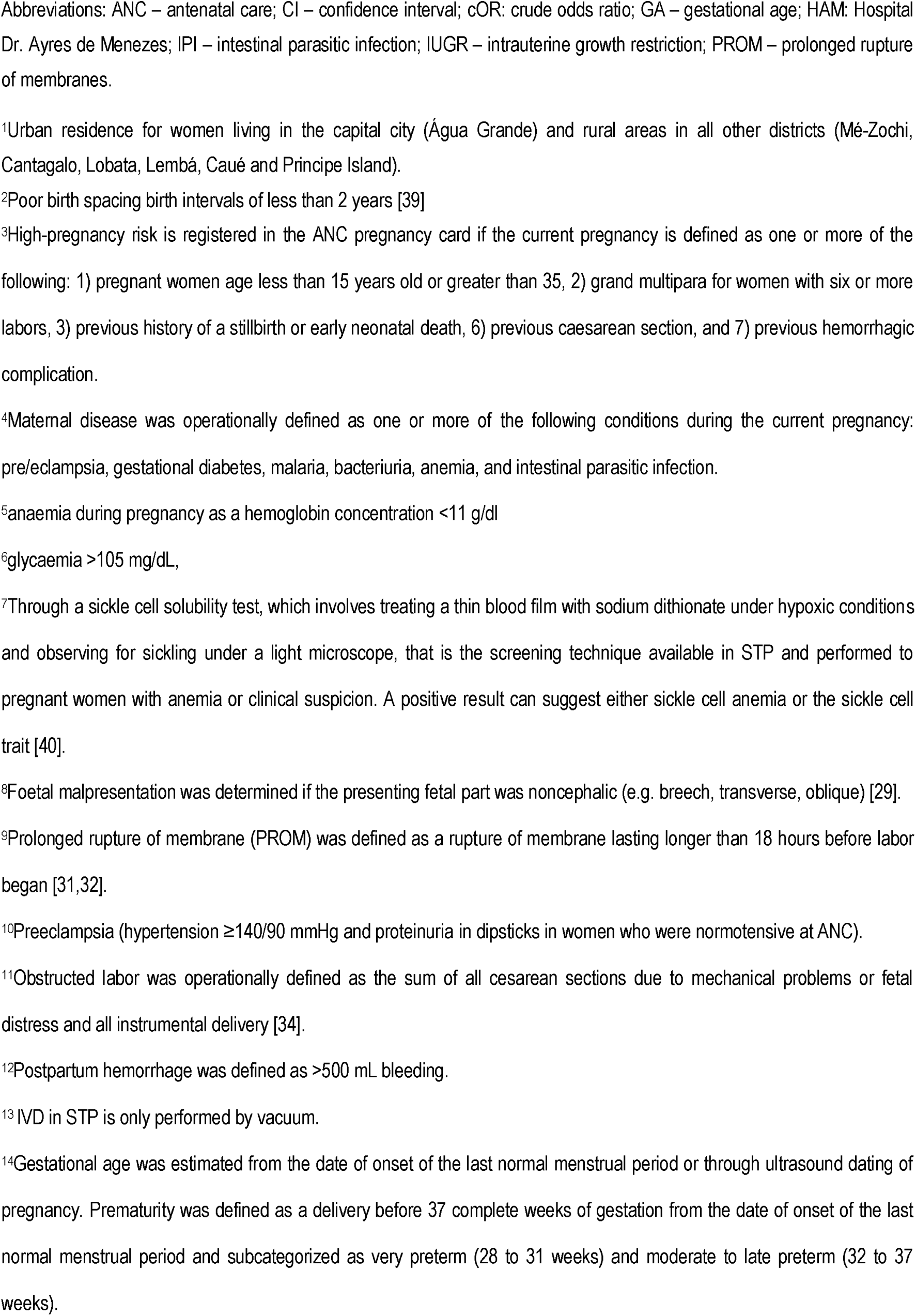
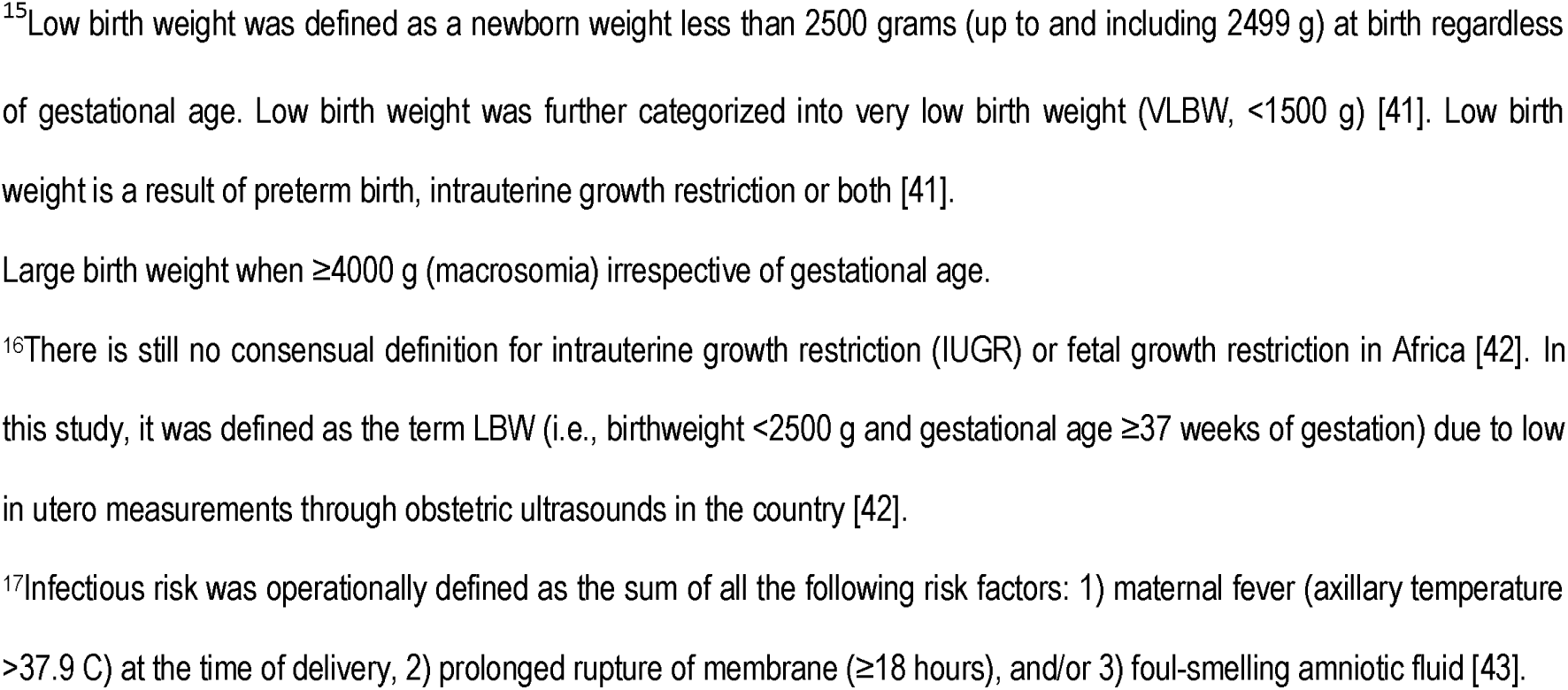
Univariable logistic regression analysis of peri-neonatal mortality among newborns admitted at HAM, Sao Tome & Principe (n= 194; death-outcome group :22 and alive-outcome group: 172)

**Table 2.** Univariable logistic regression analysis of neonatal mortality among newborns who were live births n= 178 [death-outcome group: 6 and alive-outcome group: 172]

| Variables | Categories | Total<br>n=178<sup>‡</sup> | Death-outcome<br>group<br>n=6<sup>‡</sup><br>n (%) | Alive-outcome<br>group n=172<br>n (%) | cOR | (95%) CI | p value |
| --- | --- | --- | --- | --- | --- | --- | --- |
| Neonatal resuscitation | yes | 7 (3.9) | 2 (33.3) | 5 (2.9) | 16.700 | 2.457-113.495 | **0.004** |
|  | no | 171 (96.1) | 4 (66.7) | 167 (97.1) | 1 |  |  |
| Fetal distress at birth* | yes | 26 (14.6) | 5 (83.3) | 21 (12.2) | 35.952 | 4.004-322.859 | **0.001** |
|  | no | 152 (85.4) | 1 (16.7) | 151 (87.8) | 1 |  |  |
| Birth asphyxia** | yes | 10 (5.6) | 4 (66.7) | 6 (3.5) | 55.333 | 8.421-363.600 | **<0.001** |
|  | no | 168 (94.4) | 2 (33.3) | 166 (96.5) | 1 |  |  |
| Admission at NCU | yes | 15 (8.4) | 3 (50) | 12 (7) | 13.333 | 2.425-73.310 | **0.003** |
|  | no | 163 (91.6) | 3 (50) | 160 (93) | 1 |  |  |

This is the Table 2 legend.
Abbreviations: CI – confidence interval; cOR: crude odds ratio; NCU – neonatal care unit.
<sup>‡</sup> stillbirths excluded
\*Fetal distress was defined as a low Apgar score <7 at the first minute of life (score from 0 to <7) [24,44].
\*\*Birth asphyxia was defined as a low Apgar score <7 at the fifth minute of life (score from 0 to <7) [24,44].

There were no maternal deaths in this study, with a total of 2 maternal near-misses occurring in the stillbirths’ motherś death-outcome group numbers 5 and 11 that needed hysterectomy intervention due to atonic uterus with major obstetric hemorrhage.

### Factors associated with perinatal and neonatal mortality

Binary logistic regression analysis was performed to assess the association of peri-neonatal mortality with several characteristics (Table 1 and Table 2). Crude analysis showed that meconium-stained amniotic fluid, prolonged rupture of membranes, transfer from another unit, and instrumental vaginal delivery were significantly associated with peri-neonatal mortality.

#### Sociodemographic factors

A maternal secondary education level (cOR 0.162, 95% CI 0.46–0.566, p=0.004) and a babýs father secondary education level (cOR 0.140, 95% CI 0.029–0.67, p=0.014) were found to be protective factors for peri-neonatal mortality.

#### Antepartum factors

The odds of peri-neonatal mortality were three times higher among mothers classified as having a high-pregnancy risk (cOR 2.91, 95% CI 1.18–7.22, p=0.021). Mothers without hemoglobin test during the ANC follow-up had a higher risk of having a peri-neonatal death (cOR 5.68, 95% CI 1.89–17.05, p=0.002) than those without anemia.

#### Health facility-related factors

The odds of peri-neonatal mortality were six times higher among mothers transferred from another unit compared to those directly admitted at HAM maternity (cOR 6.08, 95% CI 1.95– 18.90, p=0.002).

#### Intrapartum factors

PROM as well as meconium-stained amniotic fluid were other intrapartum factors, with the odds of mortality being almost five times higher for newborns whose mothers had a PROM (cOR 4.84, 95% CI 1.47–15.93, p=0.009), and four times higher for those with a meconium- stained amniotic fluid (cOR 4.38, 95% CI 1.74–10.98, p=0.002).

For the mode of delivery, having an instrumental assisted delivery was associated with an eightfold higher risk of perinatal and neonatal mortality (cOR 8.90, 95% CI 1.68–47.21, p=0.010).

#### Newborns’ factors

Newborns with intrauterine growth restriction had a twenty-one-fold higher risk of mortality (cOR 21.13, 95% CI 4.82-92.59, p<0.001), and newborns with an infectious risk had almost sevenfold higher odds of dying (cOR 6.61, 95% CI 2.57-16.98, p<0.001). Regarding newborn characteristics, female neonates (cOR 0.38, 95% CI 0.14-1.00, p=0.051) and birth weight greater than 2500 g and lower than 3999 g (cOR 0.017, 95% CI 0.002–0.162, p<0.001), and greater than 4000 g (cOR 0.028, 95% CI 0.001-0.564, p=0.020) were protective factors against peri-neonatal mortality.

Postpartum characteristics were only assessed for a total of six death-outcome group since sixteen stillbirths were not further included for analysis. Performance of neonatal resuscitation, fetal distress at birth (APGAR score at first-minute inferior to seven), birth asphyxia, and admission to the neonatal care unit were all related to an increased risk for neonatal mortality (Table 2).

### Multivariable model

Considering the small size of the outcome group (n=22), only one significant variable remained in the multivariable model. Newborns whose mothers had meconium-stained amniotic fluid were at a higher risk of peri-neonatal death.

## Discussion

In this prospective cohort study, the main aim was to define risk factors, the most vulnerable period for neonatal death - partum, early or late neonatal period - and to identify where they die - in the maternity, at home or in the pediatric ward - as we were able to follow them up until their 28^th^ day of life. In this study, we found that 90% died in HAM maternity before the 7^th^ day of life, that is, during the perinatal period, with mainly being stillborn (73%). The magnitude of stillbirths observed in this study (3%) is in line with the studies conducted in Nigeria (4.8%) [45] and Tanzania (3.5%) [46] and lower compared with studies from Ethiopia (6.7%) [47]. Most stillbirths occurred during intrapartum, as also identified in LMICs similar to STP, and most could have been prevented [48,49].

In this study, intrapartum characteristics, such as pregnant women with meconium-stained amniotic fluid, prolonged rupture of membranes, and instrumental vaginal delivery, were the main factors significantly associated with peri-neonatal death. Health facility-related factors, as pregnant women transferred from another unit were also at a significant risk.

High-pregnancy risk score notification was the only antepartum factor significantly associated with peri-neonatal mortality. For newborns, experiencing fetal distress at birth, needing resuscitation maneuvers, having birth asphyxia and admission to the neonatal unit were identified as high-risk factors for death. Being a female newborn and having a birth weight greater than 2500 grams were found to be protective factors.

Odds of experiencing death were identified as four times higher among newborns from mothers with meconium-stained amniotic fluid. This finding is consistent with studies from Ethiopia [6,50] and Yemen [12] that also reported higher rates of fetal death due to intrauterine passage of meconium into amniotic fluid and a fivefold increase in perinatal mortality compared with low-risk patients with clear amniotic fluid [51]. Meconium-stained amniotic fluid is associated with fetal distress since the fetus, in response, inhales the meconium, which in turn leads to airway obstruction, surfactant dysfunction and pneumonitis, which leads to loss of the fetus [6].

PROM was also a significant factor associated with neonatal mortality in this study, showing fivefold higher odds for the death-outcome group. This association was also identified in other studies from low-resource constrained countries reporting one-third of stillbirths occurring during labor as a result of prolonged labor or obstructed labor not attended to promptly [35,36,38,52,53]. The reason for this might be because PROM is a risk factor for early-onset neonatal sepsis as well as a high risk of fetal distress, respiratory distress syndrome, intraventricular hemorrhage, and death [31,32,54]. In this study we found an overall prevalence of PROM of 8.1%, which is in accordance with the global incidence that ranges from approximately 5% to 10% of all deliveries in the world [54,55].

Lack of emergency obstetric care (EmOC) is well-known to increase the risk of neonatal mortality, as laboring mothers with complications cannot immediately receive appropriate health services, such as access to a cesarean section [36]. In this study, mothers who needed to be transferred from another health unit for delivery at HAM had a sixfold higher risk of having a neonatal death outcome than mothers directly admitted at HAM. Half the mothers who were transferred from the death-outcome group had to do a 60 -kilometer journey that took approximately two to three hours until reaching HAM maternity in the capital city. Distance from a health facility with EmOC and the considerable travel times are well-known barriers and influential factors of birth adverse outcomes also found in other studies in sub-Saharan Africa [56,57].

In this study, the overall rate of 3.1% of instrumental assisted vaginal birth deliveries was found to be lower compared to 17% of births by cesarean section and 79.9% of normal vaginal deliveries. Instrumental-assisted deliveries are an effective intervention for deliveries complicated by prolonged labor or fetal distress but are also related to adverse outcomes since fetal distress or cephalopelvic disproportion are more frequently the motive to use this technique. Therefore, the ninefold higher risk of death outcome found in this study, similar to other researchers reporting [13,58,59], relates to late appropriate intrapartum care intervention with subsequently higher rates of fetal distress and risk of an intrapartum stillbirth.

Antenatal care health services in STP follow a high-risk stratification for each pregnancy and, in this study, this notification of a high-risk pregnancy in the pregnant women antenatal pregnancy card was significantly associated with a threefold higher risk for fetal and neonatal death than alive-outcome group. High-pregnancy risk was operationally defined as one or more of the following according to STP national practice, namely, age as inferior to 15 years old or superior to 35, grand multipara, previous history of a stillbirth or early neonatal death, previous cesarean section, and previous hemorrhagic complication. This is consistent with other studies that reported higher rates of perinatal and neonatal mortality with extreme maternal ages and previous adverse obstetric history (grand multipara, previous stillbirth, or early neonatal death) [1,12]. However, in this study, we could not find a statistically significant difference between groups when each of these variables were independently analyzed.

Regarding the newborns’ characteristics, the overall sex ratio at birth was 52.6% for males and 47.4% for females. The death rate was higher in male neonates (72%) than in the female death-outcome group (27.3%), with female sex being a protective factor. Sex variations are frequently reported, with a highly consistent pattern of excess male mortality across different populations and income groups [60]. Male infants are more vulnerable to fetal and early neonatal death, with most studies reporting no gender difference in mortality after 7 days of age [60]. This sex difference may be explained by a sex-specific difference in the growth and function of male and female placentae, making boys more vulnerable to different adverse outcomes [60,62].

Birth weights of more than 2500 g were protective factors for death outcome in this study. This finding is comparable with different research on other African countries that associates a birth weight of less than 2500 g (LBW) with higher rates of stillbirth and neonatal death [1]. Additionally, the odds of experiencing death were 21 times higher for babies with IUGR or fetal growth restriction defined, in this study, as a term LBW (birthweight <2500 g and gestational age ≥37 weeks) [42]. Low birth weight is a worldwide recognized important multifaceted public health problem since LBW infants are 20 times more likely to develop complications and die than normal weight infants [6,63].

We found that the odds of death were six times higher for newborns with maternal infectious risk. This finding is in agreement with previous studies conducted in SSA, as neonatal infectious risk is linked to amniotic fluid contamination that can cause intrapartum fetal infection or early postpartum neonatal infection with sepsis complicated with septic shock and multiple organ dysfunction, in which both are the most common causes of death in the perinatal period [35,43]. Infection is an important cause of stillbirth and neonatal death in LMICs, although there is a lack of overall information regarding the organisms involved, the types of transmission, and the mechanisms of death in these constrained countries [64]. In Sao Tome & Principe, malaria and syphilis are no longer an infection-burden and cause of stillbirth or neonatal death; therefore, other bacterial and viral maternal infections should be linked to stillbirth and ENND [64]. In STP, there are no means to establish a neonatal infection through blood cultures; that is, neonatal infection is based only on clinical signs with no bacteriological documentation. Additionally, nothing is known about the rate of vertical transmission of Streptococcus Group B (GBS) from colonized mothers at birth in the country. Therefore, there is a current gap in the knowledge, capability of making diagnosis and prevention and treatment of neonatal infections in STP. For instance, a study from Ethiopia [64] detected that the rate of vertical transmission of GBS from colonized mothers at birth was as high as 45.02% due to term PROM, PROM ≥18 hours before delivery and mothers having fever during labor. Detecting the risk for vertical transmission of GBS and implementing prevention methods such as adequate intrapartum administration of antibiotics are recognized as easy and affordable practices for reducing mortality due to infectious causes and should be implemented in the country along with culture techniques [43].

All four congenital major malformations detected in this study resulted in perinatal mortality, although due to the low number of newborns enrolled it was not possible to establish a statistically significant difference. Congenital malformations are known to be associated with poor outcomes for newborns, and 10–20% of stillbirths are attributed to intrinsic fetal anomalies [53]. Studies from Zimbabwe [65], Ethiopia [1] and Yemen [12] found that congenital anomalies among stillbirths were associated with a 5-fold [65], 34-fold [1] and 40- fold [12] higher risk than those among live births. Another important consideration regarding congenital anomalies is the type of malformation, since some, such as neural tube defects, are preventable and can be reduced by 30 to 50% by folic acid supplementation in pregnant women and have been estimated to cause 29% of deaths related to congenital anomalies in LMICs [4]. In this study, three out of the four major anomalies were neural tube defects, highlighting the need to enhance proper supplementation to preconceptional women in the country.

In this study, we were not able to find a higher risk associated with newborn gestational age. The high rate of stillbirths found in our study as well as the low number of preterm babies enrolled (5.6%) explain the lack of association. Stillbirths are among the most common pregnancy-related adverse outcomes worldwide, but they differ between low- and high- income countries. In high-income countries, most stillbirths occur early in the preterm period, whereas in low resource constrained countries, most occur in term or in late preterm births, as found in our study [66]. Additionally, the neonatal mortality is higher in babies born between 37 and 38 weeks of gestation than in those born between 39 and weeks [4].

From stillbirths in this study, 69% occurred during the intrapartum period, in accordance with other studies from LMICs [5]. These intrapartum stillbirths mean that intrauterine death occurred after the onset of labor and before birth (fresh stillbirth). We can also guess that some of them could be early neonatal deaths, as there are known barriers and difficulties in these contexts to establish whether a fetus or motionless newborn is living or dead after its delivery. For instance, information on some Apgar scores in these death-outcome group was not recorded, perhaps because of lack of time, especially when the neonates had to be rushed to receive resuscitation maneuvers. Studies from SSA highlight that it is very frequent that some depressed but living fetuses with a possible heartbeat do not receive resuscitation maneuvers and are prompt classified as stillbirths [1,66]. This is supported by a systematic review of sixteen hospitals and community-based perinatal mortality studies [67,68].

In this study, newborns who needed resuscitation maneuvers were at a 16-fold higher risk of dying. Additionally, the odds of neonatal death from fetal distress at birth and birth asphyxia were 36 and 55 times higher, respectively. These results are in line with studies conducted in Cameroon [69] and Ethiopia [36] as well as published literature that suggests that lower APGAR scores are associated with severe multiorgan damage resulting in brain damage, lung dysfunction, cardiomyopathy, renal failure, hepatic failure, necrotizing enterocolitis and consequently death [70]. Previous studies [23,24] that compared outcomes between adolescent pregnant girls and older counterparts identified that adverse outcomes imputable to adolescent births were fetal distress (OR 1.94, 95% CI 1.18-3.18) and performance of neonatal resuscitation maneuvers (OR 2.4, 95% CI1.07-5.38), highlighting the risks surrounding this period among deliveries at HAM maternity unit.

Perinatal asphyxia can be caused by factors grouped according to whether they are before birth (antepartum risk actors), during birth (intrapartum risk factors), or after birth (postpartum or fetal risk factors) [44,70]. Nonetheless, the single most important predictor is, undoubtedly, the quality of intrapartum care during labor and delivery.

The most important aspect of this study for public health is that it identifies potential characteristics that predispose newborns to life-threatening conditions, which is critical to address the underlying causes and provide prompt interventions by various stakeholders in the healthcare system [71].

This study allows us to perceive how to answer our question “when, where and why do newborns die in STP?” as “when”: mostly during labor, “where”: *in uterus* and “why”: mainly due to fetal distress and intrapartum-related deaths probably due to low quality and constraints of care during labor and delivery. Ending preventable stillbirths and neonatal deaths does not necessarily require new or innovative interventions. Most modifiable risk factors identified in this study can be addressed and prevented with timely, quality care during childbirth, including ongoing intrapartum monitoring and opportune intervention in case of complications [72]. Some interventions, such as computerized cardiotocography to monitor a baby’s well-being in the womb by measuring contractions, are estimated to reduce the rate of infant deaths around the time of birth by 80% when compared with traditional cardiotocography [73].

In summary, the findings of this study will be useful to health policymakers and program developers in implementing appropriate interventions to achieve the newborn health post- 2015 Sustainable Development Goals of no more than 12 neonatal deaths per 1000 live births in Sao Tome & Principe by 2030 [74].

## Strengths and Limitations

In this study, the researcher retrieved maternal and neonatal data directly from ANC cards and maternity registers to limit recall bias. The selection of the death-outcome group and alive-outcome group was based on the records of maternal and neonatal registers; therefore, it is less likely that this study has misclassification biases both in the exposure and death- outcome group _alive-outcome group categories [75].

Regarding the limitations, this is a relatively small study aiming to identify factors associated with peri-neonatal mortality in Sao Tome & Principe with a cohort of 194 newborns that were followed up until 28th days of age, 22 died and 172 survived. Thus, this study results cannot be generalized.

The newborns lost to follow-up were mainly due to mothers who provided the wrong mobile number or who did not answer the phone call at a significantly high rate; hence, the real number of early and late neonatal deaths can be much higher than the rate we have found.

Current knowledge gaps, including those associated with local burden, bacterial etiology, and eventual difficulty of nurses and mothers in perceiving subtle clinical signs associated with neonatal infections prevented us from having a clear picture of the situation in this study [43]. Data on maternal weight gain during pregnancy, nutritional status, BMI, maternal intrapartum antibiotic prophylaxis, antenatal corticosteroid administration, and total time of labor were not available.

Another limitation is that some of the variables mentioned in the univariable model had wide confidence intervals and high odds ratios due to the low number of death-outcome group enrolled in this study. This was a barrier to conducting a multivariable model, a limitation also reported in other studies [37]. Due to the small number of neonatal death-outcome group enrolled and these wide confidence intervals, the multivariable logistic regression model could only identify one independent determinant of mortality outcome.

Nonetheless, the current study can assist Sao Tome & Principe policy makers and stakeholders in designing new policies for the country to improve maternal and neonatal health outcomes.

## Conclusions

Complications such as a high-risk pregnancy score, meconium-stained amniotic fluid, prolonged rupture of membranes, being transferred from another unit, and an instrumental- assisted vaginal delivery increased the risk of stillbirth and neonatal death between 4– and 9– fold, and 90% of all these deaths occurred in the perinatal period.

Newborns with an infectious risk, intrauterine growth restriction, fetal distress at birth, who needed resuscitation maneuvers, birth asphyxia, and those admitted to the neonatal unit had a 3- to 55-fold higher risk for dying than the alive-outcome group. Female newborn and birth weight of more than 2500 g were found to be protective factors.

Implementing an efficient referral system that directs high-risk pregnancies to the EmOC facility, equipment (continuous fetal heart monitoring capability in all district maternity wards) and personnel technical skills will improve peri-neonatal outcomes and survival in Sao Tome & Principe.

## Data Availability

All relevant data are within the manuscript and its Supporting Information files.

## Declarations

### Consent for publication

Not applicable.

### Availability of data and materials

The datasets used and/or analyzed during the current study are all available within the manuscript itself.

### Competing interests

No.

### Funding

AV was supported by the Fundação para a Ciência e Tecnologia (FCT) (https://www.fct.pt/index.phtml.pt/), grant number SFRH/BD/117037/2016. The funder had no role in study design, data collection and analysis, decision to publish or preparation of the manuscript.

## Abbreviations

ANC – antenatal care; CI – confidence interval; cOR: crude odds ratio; GA – gestational age; HAM: Hospital Dr. Ayres de Menezes; IPI – intestinal parasitic infection; IUGR – intrauterine growth restriction; PROM – prolonged rupture of membranes; NCU – Neonatal Care Unit

## Authors’ contributions

AV, MCM and FP carried out the conception and design of the study. AV was responsible for field activities, data collection, and wrote the manuscript. MCM and FP critically evaluated and made progressive suggestions throughout the study and revised the manuscript. MA and ALP performed statistical analysis and reviewed the manuscript. SS and NB were involved in the study design at the country level. All the authors read and approved the final draft of the manuscript.

## Acknowledgments

A special remark for the late Professor João Luís Baptista MD PhD - AV research cosupervisor - a great man who was a thinker and a fighter for Africás improvement of public health. We are indebted to all the women who participated in the study. The authors would like to thank Elizabeth Carvalho and the 1) medical team and nurses of Hospital Dr. Ayres de Menezes Maternity unit for their support, especially to the chief-nurse Paulina Oliveira; and 2) Ana Sequeira, Rita Coelho, Ana Margalha, Ana Castro, Alexandra Coelho, and Inês Gomes for field support. We would like to acknowledge Instituto Camões, I.P. for logistic assistance in STP.

## Authors’ Information

AV is a Portuguese pediatrician with 14 years of short- and long- missions in Sao Tome & Principe, with a postgraduation in Tropical Medicine from the London Hospital of Tropical Medicine through the first East-African Course in Uganda and Tanzania and is about to finish her PhD thesis on neonatal mortality and morbidity in STP.

## Notes

### Competing Interest Statement

The authors have declared no competing interest.

### Author Declarations

The Ministry of Health of Sao Tome & Principe and the main board of the Hospital Dr. Ayres de Menezes are dedicated ethics oversight bodies, and both approved this study

### Summary of Updates

Abstract, Results and Discussion sections were updated to clarify. Supplemental files updated.

