## Additional File 1 for "Factors associated with peri-neonatal mortality in Sao Tome & Principe: a prospective cohort study"

**Additional file 1** Death-outcome group**:** characteristics of the stillbirths (n=16)

| Stillbirth number outcome group | Intrapartum stillbirths | Gestational age range^ɫ^ | sex | Birth weight | Major malformation | Pregnant Transferred | Reason for transferred | PROM | Meconium | Eclampsia | Atonic uterus | Birth attendant | Dystocia |
| --- | --- | --- | --- | --- | --- | --- | --- | --- | --- | --- | --- | --- | --- |
| 1 | no | 4 | male | 3100 | yes* | yes^1^ | no fetus heartbeat | - | yes | - | - | midwife | no |
| 2 | no | 1 | male | 1200 | - | yes^1^ | no fetus heartbeat | - | - | - | - | midwife | no |
| 3 | no | 4 | male | 2000 | - | yes^2^ | eclampsia | - | yes | yes | - | midwife | no |
| 4 | yes | 4 | male | 1900 | - | - | - | - | yes | - | - | midwife | no |
| 5 | yes | 4 | male | 2900 | - | - |  | - | - | - | yes | obstetrician | yes |
| 6 | no | 2 | female | 980 | - | yes^3^ | no fetus heartbeat | - | - | - | - | midwife | no |
| 7 | yes | 3 | female | 1750 | - | - | - | - | - | - | yes | obstetrician | yes |
| 8 | yes | 4 | male | 3300 | - | - | - | - | yes | - | - | midwife | no |
| 9 | yes | 3 | male | 1600 | - | - | - | yes | - | - | - | midwife | no |
| 10 | yes | 4 | male | 3000 | - | - | - | yes | - | - | - | obstetrician | yes |
| 11 | yes | 4 | female | 1900 | - | - | - | - | - | yes | - | midwife | no |
| 12 | no | 4 | male | 3100 | yes* | yes^1^ | no fetus heartbeat | yes | yes | - | - | midwife | no |
| 13 | yes | 4 | male | 2000 | - | - | - | - | yes | - |  | midwife | no |
| 14 | yes | 4 | male | 3500 | - | - | - | - | - | - | -- | midwife | yes |
| 15 | yes | 1 | male | 1200 | - | - | - | yes | yes | - | - | midwife | yes |
| 16 | yes | 4 | female | 3990 | - | - | - | - | yes | - | - | midwife | N0 |

This is the Table 1 legend.

*Malformation in two stillbirths: Multiple malformations in the rib cage, spina bifida, clubfoot and hydrocephalus

^1^ Caue district, ^2^ Cantagalo, ^3^ Lobata

^ɫ^sub-categories of preterm birth, based on gestational age: 1=extremely preterm (less than 28 weeks); 2=very preterm (28 to 31 weeks); 3=moderate to late preterm (32 to 36 weeks); 4=term newborns (37 to 41) and 5= posterm newborn (>42)
