## Additional File 2 for "Factors associated with peri-neonatal mortality in Sao Tome & Principe: a prospective cohort study"

**Additional file 2** Death-outcome group: characteristics of the six live births with a neonatal death (n=6)

| Neonatal death | Time of death (range) | Main cause of death | Sex | Birth weight | Prematurity | Major malformation | Birth asphyxia | NCU admission | PROM | Meconium | Eclampsia | Birth attendant | Dystocia |
| --- | --- | --- | --- | --- | --- | --- | --- | --- | --- | --- | --- | --- | --- |
| ^ɫ^ENND 1 | < 24h | Prematurity-related death | male | 1230 | yes | no | no | yes | yes | yes | no | home delivery | no |
| ENND 2 | < 24 h | Major malformation + Birth asphyxia | male | 3000 | no | yes* | yes | yes | no | no | no | obstetrician | yes |
| ENND 3 | Day 1-7 | Sepsis +  Birth asphyxia | male | 2200 | yes | no | yes | yes | no | yes | yes | midwife | no |
| ENND 4 | Day 1-7 | Major malformation +  Birth asphyxia | female | 3450 | no | yes* | yes | yes | no | yes | no | midwife | no |
| LNND 5 | Day 8-10 | Sepsis +  Birth asphyxia +  Brachial plexus lesion | male | 4250 | no | no | yes | yes | no | yes | no | midwife | no |
| LNND 6 | Day 10-15 | Probably late sepsis | female | 3800 | no | no | no | no | no | no | no | obstetrician | yes |

This is the Table 2 legend.

Abbreviations: ANC – antenatal care; NCU – Neonate Care Unit; PROM – prolonged rupture of membranes; yo – years old; ENND – early neonatal death.

*Major malformations identified ENND 2: hydrocephaly and ENND 4: thoracic malformation

^ɫ^Maternal characteristics: ENND 1 (mother age range 35-40 years old, G7 P6, 1 ANC visit); ENND 2 (mother age range 15-19 years old, G0 P0, 6 ANC visits); ENND 3 (mother age range 20-24 years old, G1 P0, 4 ANC visits); ENND 4 (mother age range 35-40 years old, G6 P5, 4 ANC visits); LNND 1 (mother age range 35-40 years old, G5 P3, 8 ANC visits); LNND 2 (mother age range 35-40 years old, G2 P1, 6 ANC visits).
